# A process evaluation of integrated service delivery of self-collected HPV-based cervical cancer screening using RE-AIM in the ASPIRE Mayuge pragmatic randomized trial

**DOI:** 10.1101/2023.05.17.23290046

**Authors:** Nadia Mithani, Anna Gottschlich, Beth A. Payne, Jessica Trawin, Arianne Albert, Jose Jeronimo, Sheona Mitchell-Foster, Ruth Namugosa, Priscilla Naguti, Angeli Rawat, Princess Nothemba Simelela, Joel Singer, Laurie W. Smith, Dirk van Niekerk, Jackson Orem, Carolyn Nakisige, Gina Ogilvie

## Abstract

**Background:** In many low-resourced settings, self-collected HPV-based cervical cancer screening (SCS) is being rolled out through task shifting to community health workers (CHWs). Process evaluations are needed to ensure SCS programs are effective and translate to community-based contexts.

**Methods:** The Advances in Screening and Prevention in Reproductive Cancers (ASPIRE) study in Mayuge, Uganda was a two-arm, pragmatic randomized trial comparing two SCS implementation strategies facilitated by CHWs: Door-to-door and Community health day recruitment. This adjunct study uses the RE-AIM evaluation framework to assess the Reach, Efficacy, Adoption, Implementation and Maintenance of each implementation strategy in a subpopulation using process data collected throughout the trial.

**Results:** Of the trial population (n=2019), 781 women participated in both the baseline and exit surveys (door-to-door: n=406; community health day: n=375) and are included in this analysis. Both implementation strategies demonstrated high Reach, Efficacy, Adoption, Implementation and Maintenance. Trial consent rate was high and 100% of consenting participants in both arms participated in SCS (Reach). Follow-up rates among HPV positive participants were also high in both arms (door-to-door: 84% and community health day: 74%) (Efficacy). The intervention employed 61 CHWs, 7 nurses, 3 health facilities and other local staff within the health system to implement the intervention (Adoption). The community health day arm received HPV screening results and visual inspection with acetic acid (VIA) quicker than the door-to-door arm, but reported higher dissatisfaction with wait times (Implementation). While women had knowledge of cervical cancer symptoms and prevention measures at six-months post-intervention, no one in either arm recalled that cervical cancer could be asymptomatic (Maintenance).

**Conclusion:** Both SCS implementation strategies performed well, demonstrating high Reach, Efficacy, Adoption, Implementation and Maintenance throughout participating communities. Implementing pragmatic approaches including task-shifting to CHWs can reduce health worker burden and improve screening access in low-resourced, community-based settings.

## Introduction

Cervical cancer, while both preventable and easily treatable at pre-cancer stages, remains a major public health burden for women in low and middle-income countries (LMICs). It is the fourth leading cause of cancer death for women globally, with over 90% of those deaths occurring in LMICs (1,2). The inequity of the cervical cancer burden between high-income countries and LMICs is mainly due to low rates of screening, follow-up and treatment, and unequal global distribution of the human papillomavirus (HPV) vaccine (3).

Because cervical cancer is a disease that is preventable through vaccination, and cervical pre cancer is treatable through early detection and treatment, the World Health Organization (WHO) issued a strategy for its elimination as a public health problem by the end of the century (4). Modelling suggests that elimination can be met by reaching the following WHO targets: 1) 90% of eligible girls vaccinated, 2) 70% of eligible women screened with a high-performance test, and 3) 90% of identified cervical disease treated (5).

HPV-based screening is considered a high performance test because it outperforms other methods of cervical cancer screening (6), such as cytology and visual inspection with acetic acid (VIA) (7). Furthermore, HPV-based screening can be self-collected, removing barriers including distance and cost-related barriers that hinder screening uptake (8) and improving screening participation among under-screened populations (9). Self-collected HPV-based cervical cancer screening (SCS), where women collect their own specimens for screening, also has the potential to reduce the burden on under resourced health systems and staff since it can be done at home without a health care provider (10).

While SCS has been shown to be acceptable (11) and feasible (12,13) across a range of settings, it remains unclear how best to implement it in resource limited settings to meet the WHO elimination targets. Program implementation strategies, such as task-shifting from physicians to nurses and community health workers (CHWs), have been proposed as cost-effective ways to improve reach of medical services in low-resourced settings (14). Realistic implementation strategies for self-collection programs, particularly in LMIC settings, include CHWs administering cervical cancer screening and education through door-to-door campaigns, or through community health meetings. According to the RE-AIM evaluation framework, to optimize the success of CHW-led, HPV-based cervical cancer screening programs, a program must reach its intended population, be effective in its impact, be adopted to the local context, be implemented as planned, and be maintained at the individual and institutional level (15).

The Advances in Screening and Prevention in Reproductive Cancers (ASPIRE) Mayuge trial compared attendance rates at treatment after a positive HPV test between two implementation strategies for SCS: Door-to-door recruitment and Community health day recruitment. To determine how these strategies were effectively implemented to achieve the outcome, and how they were received by participants within the context of a rural, community-based setting in Uganda, a process evaluation was conducted informed by the RE-AIM framework (15). The process evaluation aimed to assess the Reach, Efficacy, Adoption, Implementation and Maintenance (RE-AIM) of each implementation strategy from the perspective of the participants, as well as of the implementation process itself to understand the potential of the strategies to improve screening and treatment coverage.

## Materials and Methods

### Study design, setting and population

This is a process evaluation adjunct study of the ASPIRE Mayuge trial (16,17). ASPIRE Mayuge was a pragmatic cluster-randomized trial conducted in the Mayuge district of Eastern Uganda. Mayuge district is a primarily rural area, and has a population of approximately 480,000 people (18) Cervical cancer screening coverage in the district prior to the trial was reported at 4.8% (19) ASPIRE Mayuge compared the effectiveness of two implementation strategies of integrated SCS: Door-to-door recruitment versus Community health day recruitment. The primary endpoint, which has been previously reported (17), was attendance at treatment for HPV positive participants. All study activities, including participant recruitment and data collection, occurred between August 2019 and July 2021 and were performed within the context of the existing local healthcare system. Recruitment and participation were facilitated by trained CHWs, testing of samples was performed using the GeneXpert molecular diagnostic testing system (20) already in use within the Uganda healthcare system for HIV and tuberculosis testing, and follow-up referrals were sent to local clinics for VIA and treatment by trained nurses. Women age 25-49 years with no previous hysterectomy or treatment of cervical cancer or pre-cancer (in line with the Uganda Ministry of Health cervical cancer screening recommendations (21)) were recruited from 31 villages that were randomized to one of the two implementation strategies.

For the process evaluation, process data collected throughout the trial, including enrolment logs, clinic records, and two time-independent surveys implemented at different time points during the trial were analysed. The first survey assessed screening history, health centre access and preferences for integrated service delivery prior to trial enrolment, and the second assessed patient experience and knowledge retention of cervical cancer and screening after participation in one of the two trial-run SCS programs.

### Survey design

The ASPIRE Mayuge trial conducted a baseline survey among all participants and an exit survey among a random sample of participants conducted six months after recruitment. The baseline survey was implemented by CHWs and the exit survey was implemented by research assistants in either Lusoga or English (based on language preference of the participant). The baseline survey incorporated the Core Plus Module 1 (CPLUS1) from the Improving Data for Decision Making in Global Cervical Cancer Programs Toolkit – Part 2 (IDCCP) (22), with modifications made in collaboration with local partners to capture location-specific sociodemographic characteristics, screening history and preferences on integrated service delivery. The exit survey captured patient self-reported trial experience and knowledge retention. It was informed by the IDCCP CPLUS1 module, as well as the Organization for Economic Co-operation and Development’s (OECD) patient-reported experience measures (PREMs) survey tools (23). Additionally, knowledge retention items were taken from Mukama et. al.’s survey tool used previously in Eastern Uganda to assess women’s cervical cancer knowledge. The baseline survey items were assessed at baseline, before participants were provided with any information about cervical cancer by the study team (24). As knowledge was not assessed in the baseline survey, we use questions previously asked in similar populations as comparators. Once participants completed the baseline survey, the CHW provided them with education on cervical cancer and screening based on a WHO toolkit adopted by the Ugandan health system for all cervical cancer screening programs.

The baseline survey included questions on knowledge of cervical cancer, screening history, reason for past screen and preference for SCS at the health centre. Responses were framed as yes/no or were open-ended, with prompts for the CHWs if required. In the exit survey, patient experience questions included, motivators to participating in SCS, time to receive HPV test results and if the wait time was a problem, result of the HPV test, follow-up attendance, motivators to attending follow-up VIA and time spent waiting for VIA on the appointment day. Responses to patient experience questions were either framed as yes/no or were open-ended, with prompts for the research assistants if required. Knowledge questions included, knowledge of the recommended age for HPV vaccination and initial screening, recommended screening frequency, cervical cancer preventive measures and symptoms, and knowledge of tests that can detect cervical cancer.

### Data collection procedures and statistical analyses

In the door-to-door arm, a log was created to track the number of women approached for participation, as well as those who consented to participate. In the community health day arm, a specified number of tickets were distributed to eligible women in each community that allowed them to attend the Community health day. After recruitment, participants in both arms completed the baseline survey and were offered SCS. Samples were tested at a local lab and CHWs delivered results to participants in the same format as they were recruited (either door-to-door or at a community health day). CHWs referred HPV-positive women to treatment, which included VIA and thermal ablation, at a local health clinic and helped them schedule the appointment.

Approximately six months after recruitment, research assistants visited the villages and recontacted a subset of the study population to participate in the exit survey assessing participant experience and knowledge retention. Both of the baseline and exit surveys were conducted in a private location, either in the participant’s home or a portable tent at the community health day, and responses were recorded by the research assistant on paper. Responses were then entered in to the REDCap database software and data quality was assessed by the study team. To ensure confidentiality, identifying information was kept separate from the surveys, which only included a unique study identification number. Only the in-country team member (BP) had access to identifiable information during data collection to allow for return of HPV test results to the participants. After data collection was finalized, data was de-identified. This adjunct study includes only the subpopulation of the trial who completed both the baseline and exit surveys (n=781). Data was accessed for research purposes from April 2022 to March 2023.

All study participants provided written informed consent. Ethics approval was obtained from the University of British Columbia/Children’s and Women’s Health Centre of British Columbia Research Ethics Board (UBC C&W REB # H17–03332), the Uganda Cancer Institute (UCIREC REF-08-2018), and the Uganda National Council for Science and Technology (UNCST #HS 2517).

The primary outcomes of this analysis were used to quantify and compare the reach, effectiveness, adoption, implementation and maintenance that each implementation strategy had on its respective arm’s participants, using definitions from the RE-AIM evaluation framework, as well as other process evaluation literature (15,25). For this analysis, RE-AIM was operationalized as follows: (1) Reach as the consent and participation rates in the trial subpopulation; (2) Effectiveness as the proportion of the subpopulation who attended their follow-up appointment if they were HPV positive, and the concordance between actual and self reported HPV results; (3) Adoption (the integration of the intervention in to the local health care system and infrastructure) as the number of CHWs and health care workers trained to deliver the intervention, existing health system facilities providing care, as well as the subpopulations’ motivators/inhibitors for participating in SCS and attending VIA follow-up, (4) Implementation (the degree to which the intervention was delivered as intended) as the time it took to receive HPV test results after screening and the time it took to receive VIA follow-up on the day of the appointment, as well as if the wait time was a problem; and (5) Maintenance at the individual level as participant knowledge six months post-intervention of HPV vaccination and screening recommendations, cervical cancer and its symptoms, and cervical cancer prevention measures, given that screening education was a critical component of the intervention. At the policy level, maintenance was defined as the extent to which the intervention was embedded within the Uganda National Cervical Cancer Screening Guidelines (Table 1).

**Table 1.** RE-AIM framework metrics and indicators.

| **RE-AIM Metric** | **Primary Indicator(s)** | **Secondary Indicator(s)** |
| --- | --- | --- |
| Reach | Consent rates | Participation rates |
| Efficacy | Follow-up attendance | Concordance between actual and self-reported HPV results |
| Adoption | Number of CHWs trained, number of HCWs trained, and number of health facilities providing care | Motivators/inhibitors for SCS participation and VIA attendance |
| Implementation | Time to receive HPV results and VIA | If the wait time to receive HPV results and VIA was a problem |
| Maintenance | Knowledge of HPV vaccination and screening recommendations, cervical | Institutionalization of the intervention (policy level) |
|  | cancer, cervical cancer<br>symptoms and prevention<br>measures (individual level) |  |

Statistical analyses of baseline and exit survey data were conducted in R statistical software (26). Two sample t-tests and Chi-square or Fisher’s exact test were used to determine statistical significance between arms. Cohen’s kappa was used to determine concordance between actual HPV results and self-reported HPV results from the exit survey.

The study protocol for the ASPIRE Mayuge trial can be found at: Nakisige C, Trawin J, Mitchell-Foster S, Payne BA, Rawat A, Mithani N, et al. Integrated cervical cancer screening in Mayuge District Uganda (ASPIRE Mayuge): a pragmatic sequential cluster randomized trial protocol. BMC Public Health. 2020;20(142) (16).

## Results

### Baseline Demographics

This adjunct study describes outcomes in the subpopulation of the ASPIRE Mayuge trial (n=781). A total of 781 women participated in both the baseline and exit surveys (door-to-door: n=406; community health day: n=375). In both arms, the mean age of subpopulation participants was 34.4 years (Table 2). Over 80% of participants in each arm were married; 13% of door-to-door participants were single compared to 4% of community health day participants. Those in the door-to-door arm visited a health centre more frequently in the past year than the community health day arm (88% vs 75%) and door-to-door arm participants lived further, on average, from the nearest health centre (55 minutes vs 43 minutes). Approximately 3% of the subpopulation in each arm screened for cervical cancer prior to study participation, while in the trial population, 2% had previously screened (17). This subpopulation was comparable across all demographic factors to the trial population (17).

**Table 2.** Baseline demographics of the included study population, by study arm.

|  | Door-to-door |  | Community health day |  |  |
| --- | --- | --- | --- | --- | --- |
|  | N= | 406 | N= | 375 |  |
|  | N | % | N | % | P |
| Age (mean, SD) | 34.4 | 7.4 | 34.4 | 7.7 | 0.98 |
| Marital status | N = 405 |  | N = 375 |  | **<0.001** |
| Married | 336 | 83.0 | 337 | 89.9 |  |
| Separated/Divorced | 5 | 1.2 | 18 | 4.8 |  |
| Single | 53 | 13.1 | 14 | 3.7 |  |
| Widowed | 11 | 2.7 | 6 | 1.6 |  |
| Education | N = 405 |  | N = 375 |  | 0.65 |
| None | 47 | 11.6 | 45 | 12.0 |  |
| Primary | 226 | 55.8 | 204 | 54.4 |  |
| Secondary | 108 | 26.7 | 110 | 29.3 |  |
| Tertiary | 24 | 5.9 | 16 | 4.3 |  |
| Pregnancies (mean, SD) | 5.7 | 3.0 | 5.6 | 2.9 | 0.46 |
| Age at first birth (mean, SD) | 18.1 | 3.1 | 18 | 3.2 | 0.63 |
| Number of living children (mean, SD) | 4.8 | 2.5 | 4.8 | 2.5 | 0.73 |
| Visited a health centre in the past year | 355 | 87.7 | 280 | 74.7 | **<0.001** |
| Time from home to health centre, minutes (mean, SD) | 55.4 | 54.1 | 43.4 | 35.6 | **<0.001** |
| Ever screened for cervical cancer | 12 | 3.0 | 11 | 2.9 | 1 |
| Participated in self-collection CCS | 406 | 100.0 | 375 | 100.0 | 1 |

At baseline in the door-to-door arm, 96% (n=389) of participants had heard of cervical cancer, compared to 88% (n=329) in the community health day arm (Table 3). Among participants with a previous screen, the main reasons for screening across both arms included, being part of a routine health exam, recommended by health care provider, and experiencing pain or other symptoms. Among participants who did not report a previous screen, the main reason for never screening in both arms was lack of knowledge of how or where to get the test. Other barriers included cost, lack of time, clinic distance, and fear or embarrassment of the procedure.

**Table 3.**
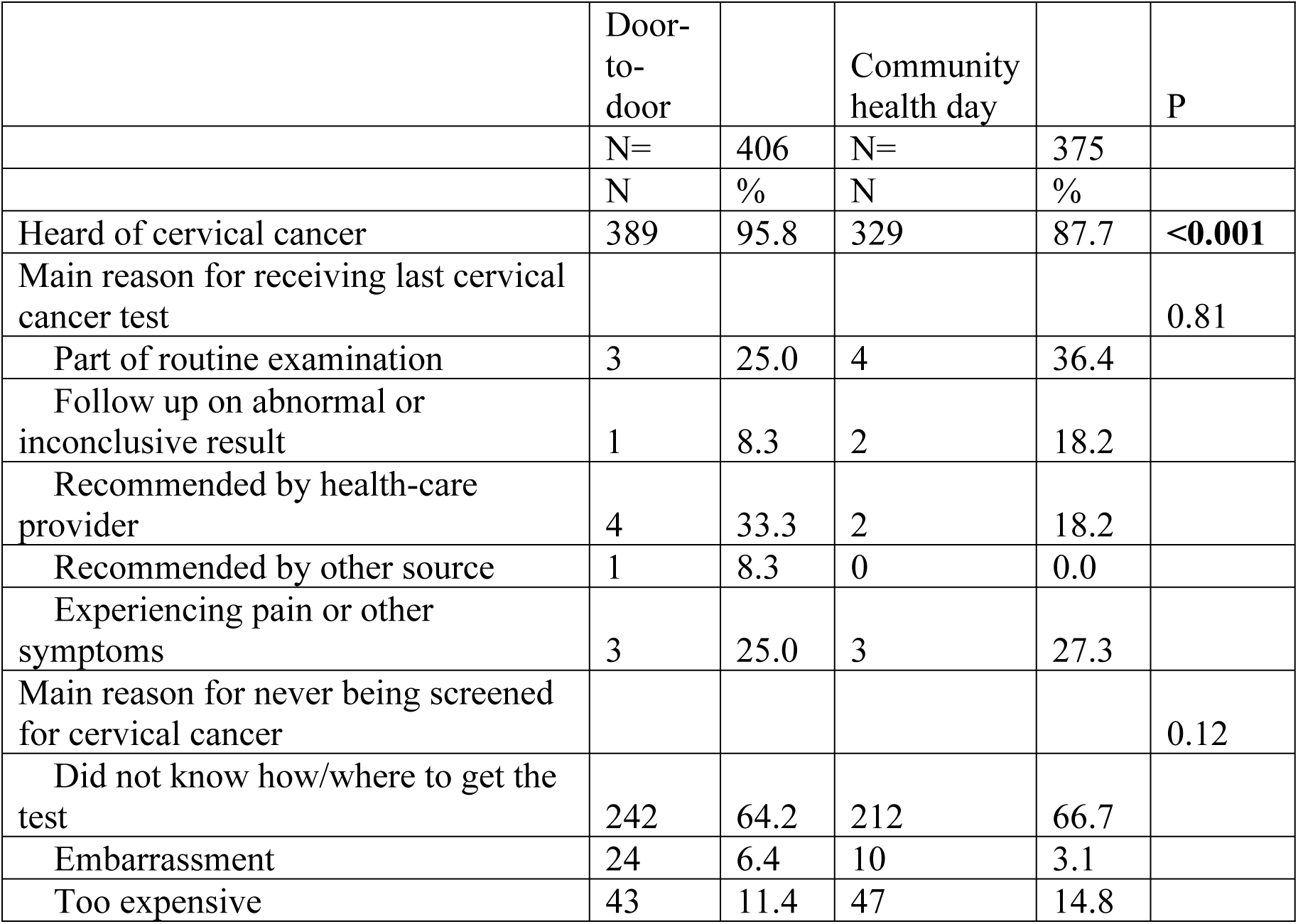
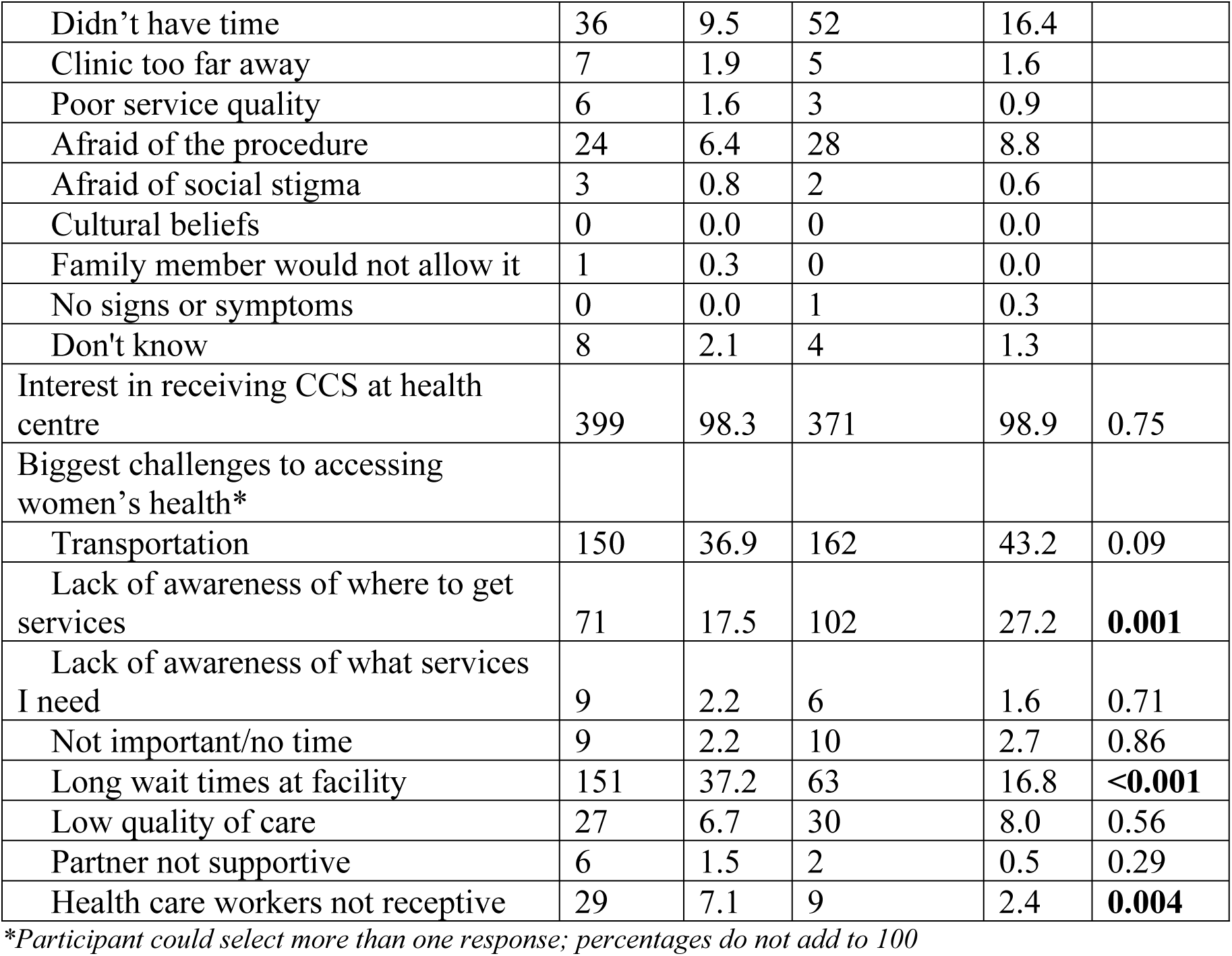
Baseline cervical cancer screening history of the included study population, by arm.

|  | Door-to-door |  | Community health day |  | P |
| --- | --- | --- | --- | --- | --- |
|  | N= | 406 | N= | 375 |  |
|  | N | % | N | % |  |
| Heard of cervical cancer | 389 | 95.8 | 329 | 87.7 | <0.001 |
| Main reason for receiving last cervical cancer test |  |  |  |  | 0.81 |
| Part of routine examination | 3 | 25.0 | 4 | 36.4 |  |
| Follow up on abnormal or inconclusive result | 1 | 8.3 | 2 | 18.2 |  |
| Recommended by health-care provider | 4 | 33.3 | 2 | 18.2 |  |
| Recommended by other source | 1 | 8.3 | 0 | 0.0 |  |
| Experiencing pain or other symptoms | 3 | 25.0 | 3 | 27.3 |  |
| Main reason for never being screened for cervical cancer |  |  |  |  | 0.12 |
| Did not know how/where to get the test | 242 | 64.2 | 212 | 66.7 |  |
| Embarrassment | 24 | 6.4 | 10 | 3.1 |  |
| Too expensive | 43 | 11.4 | 47 | 14.8 |  |
| Didn't have time | 36 | 9.5 | 52 | 16.4 |  |
| Clinic too far away | 7 | 1.9 | 5 | 1.6 |  |
| Poor service quality | 6 | 1.6 | 3 | 0.9 |  |
| Afraid of the procedure | 24 | 6.4 | 28 | 8.8 |  |
| Afraid of social stigma | 3 | 0.8 | 2 | 0.6 |  |
| Cultural beliefs | 0 | 0.0 | 0 | 0.0 |  |
| Family member would not allow it | 1 | 0.3 | 0 | 0.0 |  |
| No signs or symptoms | 0 | 0.0 | 1 | 0.3 |  |
| Don't know | 8 | 2.1 | 4 | 1.3 |  |
| Interest in receiving CCS at health centre | 399 | 98.3 | 371 | 98.9 | 0.75 |
| Biggest challenges to accessing women's health* |  |  |  |  |  |
| Transportation | 150 | 36.9 | 162 | 43.2 | 0.09 |
| Lack of awareness of where to get services | 71 | 17.5 | 102 | 27.2 | **0.001** |
| Lack of awareness of what services I need | 9 | 2.2 | 6 | 1.6 | 0.71 |
| Not important/no time | 9 | 2.2 | 10 | 2.7 | 0.86 |
| Long wait times at facility | 151 | 37.2 | 63 | 16.8 | **<0.001** |
| Low quality of care | 27 | 6.7 | 30 | 8.0 | 0.56 |
| Partner not supportive | 6 | 1.5 | 2 | 0.5 | 0.29 |
| Health care workers not receptive | 29 | 7.1 | 9 | 2.4 | **0.004** |

\*Participant could select more than one response; percentages do not add to 100

Over 98% of participants in each arm were interested in receiving cervical cancer screening at a local health centre. The biggest reported challenge to accessing women’s health services in both arms was transportation (door-to-door: 37%, n=150; community health day: 43%, n=162). For other barriers, the reported frequency differed by arm. A higher percentage of women in the door-to-door arm (37%, n=151) than in the community health day arm (17%, n=63) reported long wait times at facilities and unresponsive health care workers as challenges, while a higher percentage of women in the community health day arm (27%, n=102) than the door-to-door arm (17%, n=71) reported lack of awareness of where to get services as a challenge. Few participants in either arm reported lack of awareness of services they needed, services not being important, or partner not supportive as challenges to accessing women’s health care.

## Reach – trial participation

### Primary indicator

In determining consent rate, due to a miscommunication with the CHWs in the door-to-door arm, women who declined to participate in the trial were not entered into the consent log. This error was not detected until completion of the arm. However, upon detection, qualitative interviews were conducted with each CHW to assess their perception of consent rate. CHWs reported that few women declined participation in the trial. In the community health day arm, a total of 1,200 tickets were distributed, with 964 women attending the Community health days, resulting in an 80.3% consent rate for the community health day arm.

### Secondary Indicator

Among those who consented to trial participation (n=2,019), 100% of participants in both arms consented to SCS (17). Similarly, 100% of subpopulation participants in both arms (n=781) consented to SCS (Table 2). As previously reported, nine total participants did not receive their HPV test results and were considered lost to follow-up (Door-to-door: n=5, Community health day: n=4) (17).

## Effectiveness – trial experience

### Primary indicator

Attendance at treatment follow-up by arm in the trial population has been previously reported (17). Among the HPV positive participants in the subpopulation (n=205), both arms had high rates of follow-up attendance (door-to-door: 84%, community health day: 74%) (Table 4). Rates were slightly higher in the door-to-door arm, which is consistent with the findings from the trial population (17).

**Table 4.**
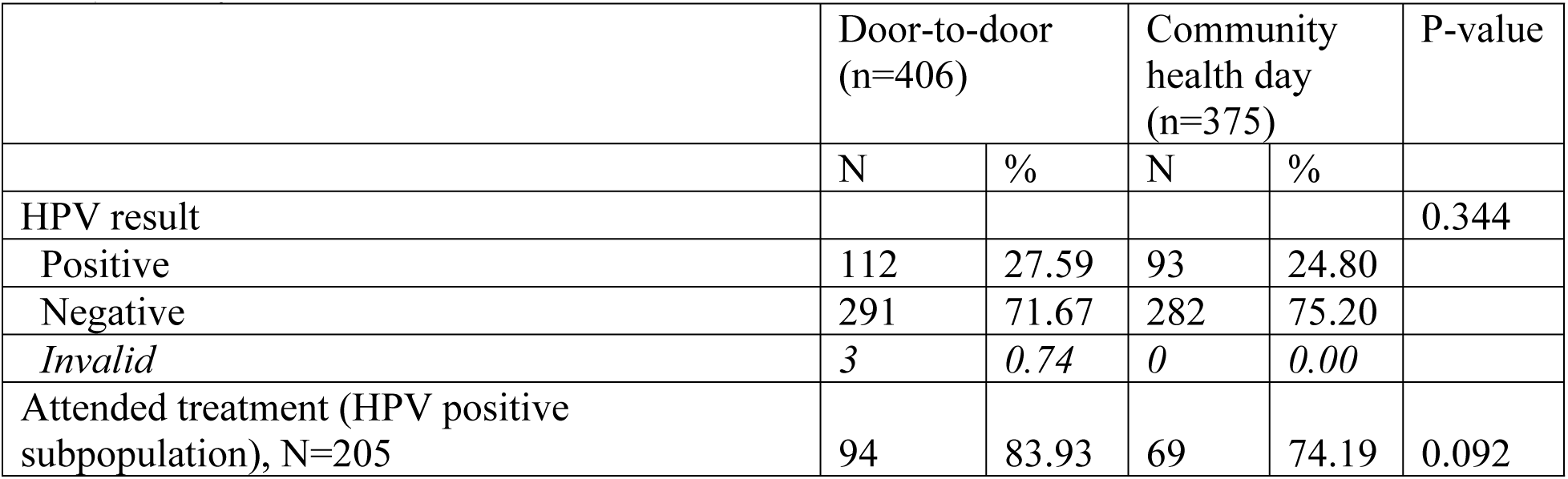
Treatment attendance among HPV positive in the included study population, by arm (Efficacy)

|  | Door-to-door<br>(n=406) |  | Community<br>health day<br>(n=375) |  | P-value |
| --- | --- | --- | --- | --- | --- |
|  | N | % | N | % |  |
| HPV result |  |  |  |  | 0.344 |
| Positive | 112 | 27.59 | 93 | 24.80 |  |
| Negative | 291 | 71.67 | 282 | 75.20 |  |
| *Invalid* | 3 | 0.74 | 0 | 0.00 |  |
| Attended treatment (HPV positive subpopulation), N=205 | 94 | 83.93 | 69 | 74.19 | 0.092 |

### Secondary Indicator

Concordance rates were determined from actual HPV results and those self-reported in the exit survey (Table 5). In the door-to-door arm, 97/111 HPV positive participants reported that they were positive (87%), compared to 89/93 in the community health day arm (96%). The negative agreement rate was similar across arms (door-to-door: 288/290, 99.3%; community health day: 281/282; 99.6%). Cohen’s kappa for the door-to-door arm was 0.88 and 0.96 for the community health day arm, both representing excellent agreement.

**Table 5.** Concordance between actual and self-reported HPV results, by arm (Efficacy)

|  |  |  |  |  |  |
| --- | --- | --- | --- | --- | --- |
| Door-to-door | Actual results |  |  |  |  |
| Self-reported | Positive | Negative | Inconclusive | Did not receive | Cohen's kappa |
| Positive | 97 | 2 | 0 | 1 | 0.88 |
| Negative | 7 | 288 | 2 | 1 |  |
| Inconclusive | 6 | 0 | 1 | 0 |  |
| Did not receive results | 1 | 0 | 0 | 0 |  |
| Community health day | Actual results |  |  |  |  |
| Self-reported | Positive | Negative | Inconclusive | Did not receive | Cohen's kappa |
| Positive | 89 | 1 | 0 | 0 | 0.96 |
| Negative | 4 | 281 | 0 | 0 |  |
| Inconclusive | 0 | 0 | 0 | 0 |  |
| Did not receive results | 0 | 0 | 0 | 0 |  |

## Adoption – trial integration

### Primary indicator

The integration of the intervention in to the local Ugandan health care system, as measured by the number of CHWs trained to deliver SCS, has been previously reported. 61 existing CHWs from the 25 villages selected for the intervention were recruited and trained to deliver SCS (17). Seven nurses already providing cervical cancer screening with VIA at the selected health centres for the intervention were given refresher training on VIA and treatment of precancerous lesions with thermal ablation. Three existing government health centres in close proximity to the 25 intervention villages provided the infrastructure for follow-up and treatment of HPV positive participants and one providing laboratory infrastructure as well. Participants with advanced lesions were referred to the Uganda Cancer Institute (UCI) for further evaluation and treatment. An established laboratory staff member and data analyst were employed for the duration of the study.

### Secondary indicators

The highest reported motivator to participating in SCS was having support from a CHW (door-to-door arm: 39%, n=159; community health day arm: 29%, n=109) (Table 6). Many door-to-door participants also reported wanting to know their HPV status (35%, n=142), while community health day participants more often reported knowing the potential health consequences for not screening (19%, n=70) and having access to SCS (12%, n=44) as motivators to SCS participation. Among HPV positive participants, the main factor motivating VIA follow-up attendance was receiving an HPV positive test result (door-to-door arm: 61%, n=58, community health day arm: 81%, n=57). CHW support was also reported as a motivator for follow-up, particularly in the door-to-door arm (17%, n=16).

**Table 6.** Reported motivators to screening and treatment at exit, by arm (Adoption)

|  | Door-to-door |  | Community health day |  | P |
| --- | --- | --- | --- | --- | --- |
|  | N= | 406 | N= | 375 |  |
|  | N | % | N | % |  |
| Main motivator to participate in SCS |  |  |  |  | **<0.001** |
| Having access to screening through self-collection in my community | 14 | 3.4 | 44 | 11.7 |  |
| Having support from a CHW | 159 | 39.2 | 109 | 29.1 |  |
| Knowing that other women in my community are being screened for cervical cancer | 15 | 3.7 | 34 | 9.1 |  |
| Knowing a women who has been diagnosed with or died from cervical cancer in the past | 15 | 3.7 | 37 | 9.9 |  |
| Knowing the potential health consequences of not screening for cervical cancer | 16 | 3.9 | 70 | 18.7 |  |
| Being provided with instructions and visuals for self-collection | 20 | 4.9 | 10 | 2.7 |  |
| Having symptoms such as pain, bleeding, or discharge | 19 | 4.7 | 26 | 6.9 |  |
| Wanted to know HPV status | 142 | 35.0 | 45 | 12.0 |  |
| Refused/Missing | 6 | 1.5 | 0 | 0.0 |  |
| Main motivator to follow-up with VIA after receiving a positive HPV test result |  |  |  |  | **0.03** |
| Receiving a positive HPV test result | 58 | 61.1 | 57 | 81.4 |  |
| Knowing the potential health consequences associated with a positive HPV test result | 4 | 4.2 | 4 | 5.7 |  |
| Having support from a CHW | 16 | 16.8 | 4 | 5.7 |  |
| Knowing a women who has been diagnosed with or died from cervical cancer in the past | 3 | 3.2 | 0 | 0.0 |  |
| Knowing that other women in my community are being screened for cervical cancer | 0 | 0.0 | 0 | 0.0 |  |
| Setting a date for when to follow-up with VIA | 1 | 1.1 | 0 | 0.0 |  |
| Being provided with instructions on how and where to follow-up with care | 0 | 0.0 | 0 | 0.0 |  |
| Wanted to confirm status | 6 | 6.3 | 3 | 4.3 |  |
| Wanted treatment | 6 | 6.3 | 1 | 1.4 |  |
| Had symptoms | 0 | 0.0 | 1 | 1.4 |  |
| Missing | 1 | 1.1 | 1 | 1.4 |  |

## Implementation – trial delivery

### Primary indicator

Community health day participants received their HPV test results quicker than door-to-door arm participants did. In the community health day arm, 38% (n=143) received their results within 6 days and 79% (n=296) within 14 days, compared to the door-to-door arm, where 7% (n=27) received results within 6 days and 67% (n=273) within 14 days (Table 7).

**Table 7.** Reported wait times for results at exit, by arm (Implementation)

|  | Door-to-door |  | Community health day |  | P |
| --- | --- | --- | --- | --- | --- |
|  | N= | 406 | N= | 375 |  |
|  | N | % | N | % |  |
| How quickly did you get your HPV test result? |  |  |  |  | **<0.001** |
| 1-6 days | 27 | 6.7 | 143 | 38.1 |  |
| 7-14 days | 246 | 60.6 | 153 | 40.8 |  |
| 15-30 days | 127 | 31.3 | 77 | 20.5 |  |
| Don't know | 6 | 1.5 | 2 | 0.5 |  |
| Was the time you waited to receive your HPV test result a problem for you? | 97 | 23.9 | 138 | 36.8 | **<0.001** |
| Follow up visits | 95 | 23.4 | 70 | 18.7 | 0.11 |
| How long did you wait for VIA on day of visit |  |  |  |  | 0.06 |
| 0-15 mins | 38 | 40.0 | 24 | 34.3 |  |
| 15-30 mins | 24 | 25.3 | 29 | 41.4 |  |
| 30-60 mins | 14 | 14.7 | 13 | 18.6 |  |
| 1-2 hours | 11 | 11.6 | 2 | 2.9 |  |
| 2-4 hours | 4 | 4.2 | 0 | 0.0 |  |
| 4-8 hours | 1 | 1.1 | 0 | 0.0 |  |
| More than 8 hours | 0 | 0.0 | 0 | 0.0 |  |
| Left before being seen | 2 | 2.1 | 2 | 2.9 |  |
| Don't know | 1 | 1.1 | 0 | 0.0 |  |
| Time problem |  |  |  |  | **0.001** |
| Yes | 9 | 9.5 | 21 | 30.0 |  |
| No | 85 | 89.5 | 49 | 70.0 |  |
| Missing | 1 | 1.1 | 0 | 0.0 |  |
| Reason time problem |  |  |  |  | 0.14 |
| Responsibilities (work, caring for children) | 1 | 11.1 | 1 | 4.8 |  |
| Lack of support from husband | 0 | 0.0 | 6 | 28.6 |  |
| Time out of day | 3 | 33.3 | 2 | 9.5 |  |
| Stress about results | 5 | 55.6 | 12 | 57.1 |  |
| Received treatment |  |  |  |  | 0.27 |
| Yes | 82 | 86.3 | 66 | 94.3 |  |
| No | 11 | 11.6 | 4 | 5.7 |  |
| Refused/Missing | 2 | 2.1 | 0 | 0.0 |  |

### Secondary indicators

However, in the door-to-door arm, 24% reported that the amount of time waited to receive HPV test results was an issue for them, compared to 37% in community health day arm. Similarly, more door-to-door arm participants spent 15 minutes or less waiting for VIA follow-up on the day of their appointment than community health day arm participants (40%, n=38 vs 34%, n=24 respectively). However, overall community health day participants spent less time waiting for VIA follow-up, with 94% (n=66) waiting one hour or less compared to 80% (n=76) of door-to-door participants. Similar to screening, wait time was more of an issue in 30% of community health day participants compared to 9% of door-to-door participants.

## Maintenance – knowledge retention after trial

### Primary Indicators

At exit, nearly 100% of included study participants in each arm reported having heard of cervical cancer (Table 8). In both arms, most participants believed the recommended vaccination age was between 6 and 15, the recommended age to start screening was between 16 and 30, and the recommended screening frequency was yearly. More women in the door-to-door arm than the community health day arm responded that early screening was a preventive measure against cervical cancer (97% vs. 80%), as well as avoiding multiple sexual partners (16% vs. 9%). Over 80% of women in both arms responded that vaccination was another preventive measure.

**Table 8.**
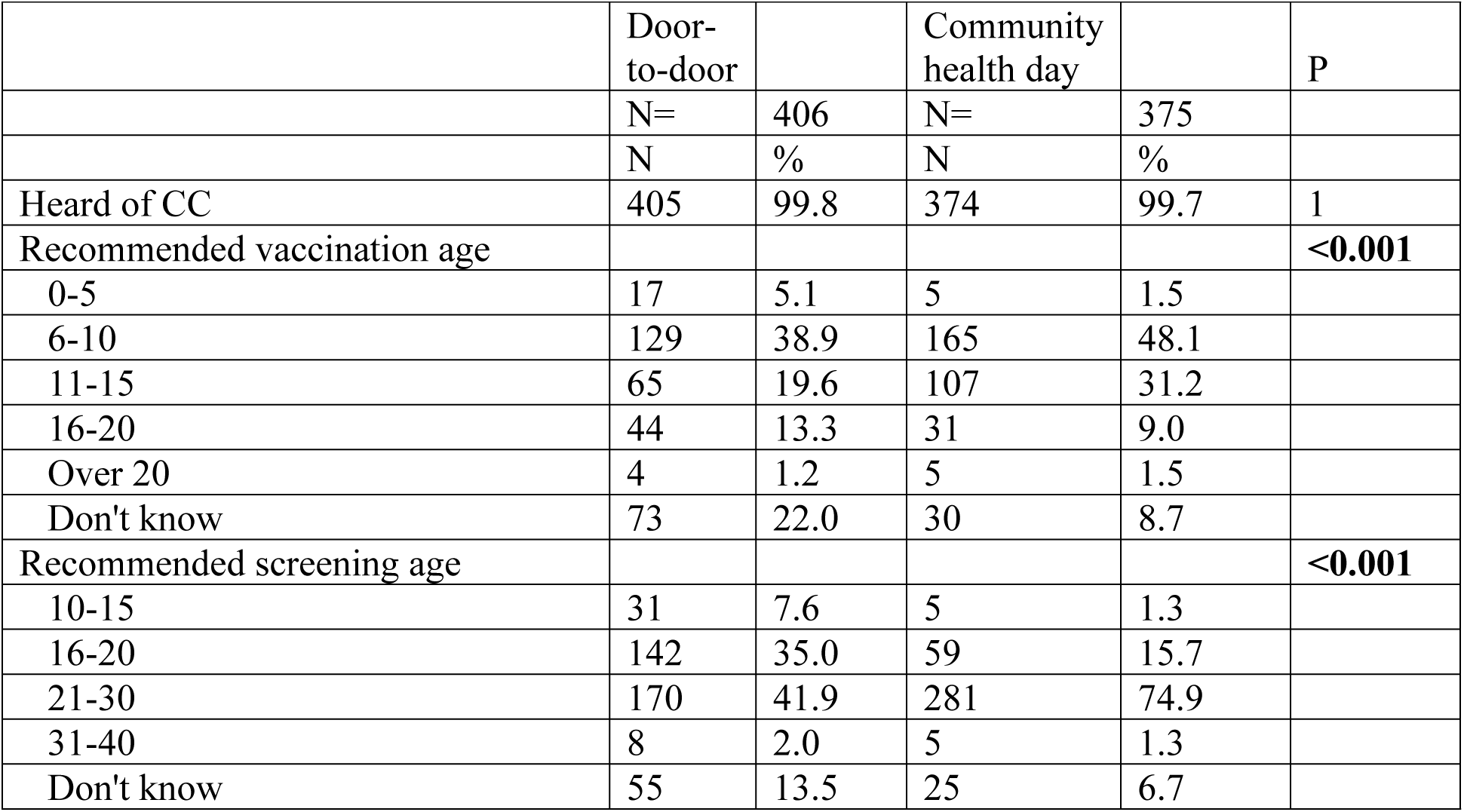
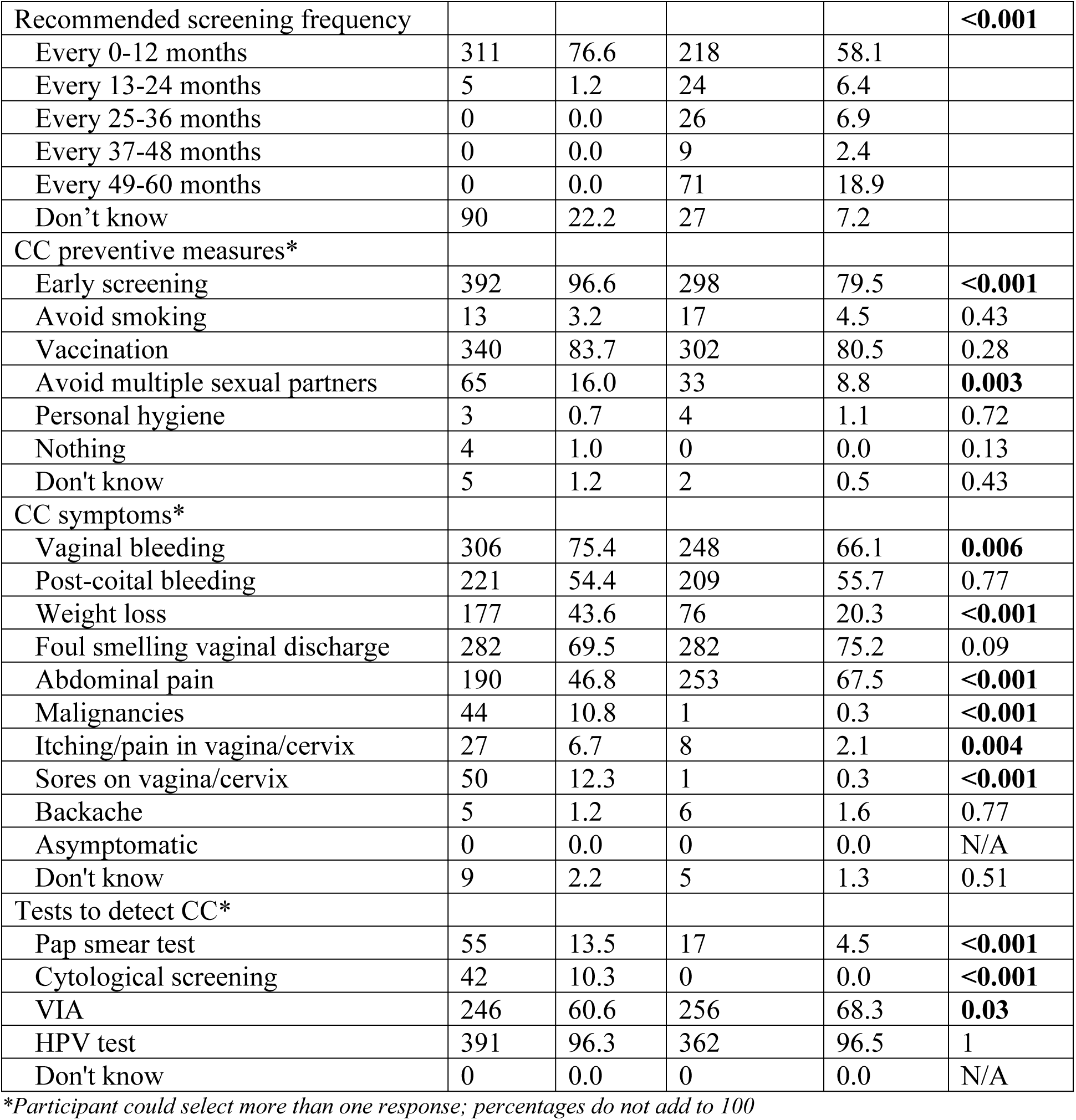
Cervical cancer knowledge at exit, by arm.

Women in both arms responded that vaginal bleeding, post-coital bleeding, weight loss, pain, and discharge were symptoms of cervical cancer; no one in either arm reported that cervical cancer could be asymptomatic. Over 96% and 60% of women in each arm reported that HPV testing and VIA could detect cervical cancer, respectively. More women in the door-to-door arm than the community health day arm had heard of Pap tests and cytology screening to detect cervical cancer.

### Secondary Indicator

This SCS intervention has already begun to inform the Uganda National Cervical Cancer Screening Guidelines and change how cervical cancer screening services are provided at the community level. Prior to the intervention, screening was offered at the health facility level only. Since the trial, SCS is beginning to be offered at the household level by CHWs. A draft version of the national screening guidelines has been developed outlining the process of screening through SCS, as well as the emphasis on follow-up and treatment (the primary outcome of the trial).

## Discussion

This process evaluation of the ASPIRE Mayuge trial found high Reach, Efficacy, Adoption, Implementation and Maintenance for both the door-to-door and community health day implementation strategies for integrated self-collected HPV-based cervical cancer screening programs. This suggests the future success of these programs in rural, community-based settings and their potential to be incorporated in to national cervical cancer prevention and control programs. Women were engaged in cervical cancer screening and follow-up, and viewed preventive care as an important women’s health issue. This adds to the main trial findings that both strategies for implementation of SCS were feasibly integrated into the existing health system through the use of CHWs and existing health infrastructure and staff, and significantly increased screening and treatment coverage (17). Many studies have shown that SCS is highly accepted (11,27–30) and performs similarly to physician-collected samples (31); this study adds to the literature by demonstrating an example of a successful, real-world implementation of a self-collection screening program to increase screening and treatment uptake in an under resourced setting.

We found that both arms achieved high reach within the community, with majority of contacted women consenting to participate in the trial. Many studies have found that women across a variety of settings are interested in participating in health promotion campaigns, particularly in cancer screening (13,27,30,32–34), as we found in this population. Furthermore, as previously reported, 100% of the trial population (and thus subpopulation) across both arms chose to participate in the self-collection screening opportunity that followed the baseline survey. Prior to enrolment, only about 3% of the subpopulation and 2% of the trial population had ever been screened for cervical cancer (17).

Additionally, around 80% of HPV positive participants in the subpopulation attended follow-up, demonstrating high efficacy of the intervention. This study was uniquely designed to allow for investigation, not only of the uptake of participation in screening programs, but also the resulting treatment coverage among those with positive screens, an issue often omitted in self-collection cervical cancer screening literature. While participants across both arms generally accurately reported their HPV results, community health day participants had higher concordance between actual and reported results.

There was a high level of adoption of both implementation strategies and their integration in to the local health system. Those offering the intervention (CHWs, nurses and staff) were all part of the local community in Mayuge district and already embedded within the established health care system. Training of existing health system staff and use of existing health facilities increases adoption of the intervention by staff, the communities and the health system. Understanding the setting and system within which an intervention is occurring is necessary for its adoption, as well as its sustainability beyond the intervention delivery (15). Notably, one of the main motivators to participating in SCS and attending VIA follow-up was CHW support. CHWs have been shown to increase screening and treatment coverage rates in LMICs, particularly with respect to facilitating linkage to follow up care (35).

Another primary motivator to participate in SCS was participants’ desire to know their HPV status. Most prior studies note that women choose to participate in SCS programs because they reduce barriers to screening in conventional programs (such as cost, travel, and embarrassment) (9,36,37). We hypothesize that this novel finding has to do with the common language used by both HIV and HPV screening and treatment programs in this setting. One study from Malawi recently found that while participants had minimal knowledge of cervical cancer, they wer knowledgeable about screening programs, likely stemming from HIV screening and prevention concepts discussed in the community (38). Common language can be useful to promote education, health behaviours and the integration of HIV and HPV screening services in LMICs (39). However, it is also important to carefully consider and effectively differentiate the implications of their diagnoses in order to promote adequate health seeking behaviour and avoid further stigmatization.

The degree to which the intervention was implemented as intended, defined by screening and follow-up wait times, was moderately high. Majority of participants in both arms received their HPV test results within two weeks and did not wait more than one hour for VIA. Community health day participants received their HPV results quicker and waited for less time for VIA than door-to-door participants; yet, community health day participants were more likely to report that wait time for both SCS and VIA was an issue. This may be in response to frustration from significant health care delays and disruptions that have been common around the world throughout the COVID-19 pandemic (40) when the community health day arm was being implemented.

The maintenance of the intervention at the individual level, defined as knowledge retention six months post-intervention, was quite high by the end of the trial. Almost all participants had heard of cervical cancer after the trial. In general, women knew that early screening and vaccination were prevention measures, that an HPV test detected cervical cancer, as well as recognized common symptoms of cervical cancer. This suggests that the information provided by CHWs during the baseline visit was retained for at least six months. However, similar to the findings in Mukama et. al, participants in both arms of our trial had less understanding of the recommended vaccine age and screening initiation age and frequency (24). Of note, regarding disease symptoms, zero participants reported that cervical cancer could be asymptomatic. This knowledge is critical to recognize the importance of screening, thus future education materials should make this more explicit.

Maintenance at the policy level was defined as the institutionalization of the intervention. Although still in draft format, the findings from this intervention have been incorporated in to the Uganda National Cervical Cancer Guidelines, particularly the use of SCS and CHWs for screening at the household and community level in rural areas. Additionally, implementation of household-based cervical cancer screening through the use of CHWs has begun since this trial was completed.

This study provides valuable new evidence from the implementation of two strategies for self collection HPV-based cervical cancer screening using data from a pragmatic randomized trial. However, the results of this study must be considered with its limitations. Although CHWs noted a low refusal rate to consenting to the study, the actual consent rate for the door-to-door arm could not be calculated. Because CHWs implemented the baseline survey, self-collection, and results dissemination, agreement bias may have influenced women’s survey responses; however, research assistants were deployed to administer the exit survey to limit this concern.

Additionally, community health day study activities were conducted during the COVID-19 global pandemic. The pandemic caused disruptions to health care around the world and may have impacted participant responses to the acceptability of the intervention. Lastly, while the protocol planned for both arms to complete the exit survey six months after baseline, due to delays, door-to-door arm participants were completed it closer to one-year post-baseline, while community health day arm participants completed it at the planned six-month interval. This time difference may be a factor in reporting differences across arms, including the possibility of recall bias for door-to-door arm participants. Regardless, across both arms, participants were highly satisfied with the quality of their respective intervention strategy. Finally, the knowledge questions were only assessed six-months post-recruitment, so we cannot investigate an increase from baseline.

The recent global strategy towards the elimination of cervical cancer as a public health burden has encouraged countries around the world to expand their respective screening programs. In our study, participants reported that the main reason they had never been screened for cervical cancer was that they did not know how or where to get the test. This finding highlights the importance of educational components integrated into self-collection HPV-based cervical cancer screening campaigns. Participants also reported that some of the biggest challenges to accessing women’s healthcare were transportation and long wait times at health facilities. Our implementation strategies reduced these barriers by bringing screening information closer to the community.

This process evaluation demonstrated that to achieve the primary outcome, the process of implementing SCS in a rural, community-based setting required a thoughtful, pragmatic approach that incorporated the setting and context of the intervention, how its implemented, and participant views on how they received the intervention (42). Both implementation strategies were viable options to improve cervical cancer control in a high-burdened setting. However, providing information and screening to under screened populations through CHWs, understanding motivators and inhibitors to screening and treatment, and bringing services closer to the community improved knowledge and access, and empowered women to improve their health, while the exact mode of delivery was perhaps secondary. National programs should focus on scaling up community-based services so that women can access the care they want and deserve. Future work will aim to investigate differences by age and setting (i.e., rural vs. urban) and compare the cost-effectiveness of each implementation strategy, using detailed costing data collected throughout the trial, to provide additional insight into the feasibility of each strategy.

## Data Availability

All data produced in the present study (restricted to nonidentifying data owing to privacy concerns) can be requested for scientific purposes only from the corresponding or senior authors, who will handle all requests.

## Acknowledgements

We would like to acknowledge the contributions of the staff at the Uganda Cancer Institute who assisted in the setup of the trial, as well as local lab managers, hub riders, Kigandalo Health Centre hospital administrations, and the CHWs.

